# The role of corticosteroids in the management of critically ill patients with coronavirus disease 2019 (COVID-19): A meta-analysis

**DOI:** 10.1101/2020.04.17.20069773

**Authors:** Kalyan Kumar Gangopadhyay, Jagat J Mukherjee, Binayak Sinha, Samit Ghosal

## Abstract

**Objective:** There are no controlled studies on the role of systemic corticosteroids (CS) in patients with coronavirus disease 2019 (COVID-19). In the absence of high-quality evidence, understandably the recommendations from various organizations are cautious. Several randomized controlled trials are underway but shall take time to conclude. We therefore undertook a meta-analysis to ascertain the role of CS in the management of critically ill patients with COVID-19.

**Data Sources:** Electronic databases, including Pubmed, Cochrane library and Embase, were searched, using the keywords of interest and the PICO search technique, from inception to 12th April 2020.

**Study Selection:** Studies highlighting the use of CS in coronavirus infection with severe acute respiratory syndrome (SARS), Middle East Respiratory Syndrome (MERS) and COVID-19 were selected based on pre-determined inclusion criteria.

**Data extraction:** Data was extracted into an excel sheet and transferred to comprehensive meta-analysis software version 3, Biostat Inc., Englewood, NJ, USA, for analysis.

**Data synthesis:** Five studies with SARS-CoV-2 infection were included in the meta-analysis. The rate ratio (RR) for mortality in patients with SARS-CoV-2 infection was 1.26 (95% CI: 0.96-1.65, I^2^: 74.46), indicating lack of benefit of CS therapy on mortality in critically ill patients with COVID-19. The RR for mortality on analysis of the three studies that particularly reported on patients with significant pulmonary compromise secondary to SARS-CoV-2 infection was neutral (RR: 0.91, 95% CI: 0.63-1.33, I^2^: 63.38).

**Conclusions:** The use of CS in critically ill patients with COVID-19 did not improve or worsen mortality. Pending further information from controlled studies, CS can be used in critically ill patients with COVID-19 with ‘critical illness related corticosteroid insufficiency’ and moderate to severe ARDS without the risk of increased mortality.

## 1.0 Introduction

The severe acute respiratory syndrome coronavirus 2 (SARS COV2) pandemic that started in late 2019 from Wuhan district in China has created a havoc on human civilization, with poverty, destruction and death dogging every aspect of human life across the world (1). The disease, named coronavirus disease 2019 (COVID-19), is mild or asymptomatic in the majority, but causes a devastating pneumonia with bilateral lung infiltrates in some, leading to hospitalization and a high risk of acute respiratory distress syndrome (ARDS), shock, cytokine storm syndrome and death (2). In the absence of high quality evidence, experts remain divided as to the usefulness of systemic corticosteroid (CS) therapy in this situation (3).

There is no debate about the usefulness of continuing CS in stress doses in patients with COVID-19 who are on replacement steroids for adreno-cortical insufficiency, and in patients who are on long term CS therapy for chronic diseases like rheumatoid arthritis or asthma (4).

The experience with use of CS in patients with complications of pneumonia during previous coronavirus epidemics due to severe acute respiratory syndrome (SARS) and Middle East Respiratory Syndrome (MERS), was mixed (5,6). In a retrospective observational study, 151 of 309 patients with MERS, who were treated with CS, were found to be more likely to require mechanical ventilation, vasopressors, and renal replacement therapy (5). Use of CS was associated with delayed clearance of viral RNA from respiratory tract secretions (5). Similarly, a systematic review and meta-analysis of four studies documenting the use of CS in patients with SARS showed delayed clearance of viral load, increased evidence of diabetes mellitus, psychosis and avascular necrosis in patients receiving CS(6).

In view of lack of benefit, and possible harm, seen with CS therapy in patients with SARS and MERS, the World Health Organization (WHO) (7), the center for diseases and prevention (CDC) (8), and the infectious diseases society of America (9) have all recommended against the use of CS, particularly in high doses. In contrast, the Surviving Sepsis Campaign COVID-19 panel have made a weak recommendation for using CS in mechanically ventilated adults with COVID-19 (10).

In the absence of high-quality evidence, understandably the recommendations from various organizations are cautious and dichotomous. Several randomized controlled trials on use of CS in patients with COVID-19 are underway but shall take time to conclude (Supplementary table 2) (11-14). We therefore undertook a meta-analysis to ascertain the role of CS in the management of critically ill patients with COVID-19.

## 2.0 Materials and methods

An electronic database search was conducted using the Cochrane library, PubMed and Medline. Search keywords included “steroids”, “corticosteroids”, “hydrocortisone”, “prednisolone”, “dexamethasone”, “methylprednisolone”, “SARS”, “SARS-CoV”, “severe acute respiratory syndrome”, “MERS”, “MERS-CoV”, “middle east respiratory syndrome”, “COVID19”, “coronavirus disease 2019”, “SARS-CoV-2”, “severe acute respiratory syndrome coronavirus 2”, “mortality”, “death”, “complications”, “acute respiratory distress syndrome” and “viral pneumonia”. As a part of advanced screening references of identified citations were also screened for any additional information missed out in the screening process. [Figure 1]

**Figure 1:**
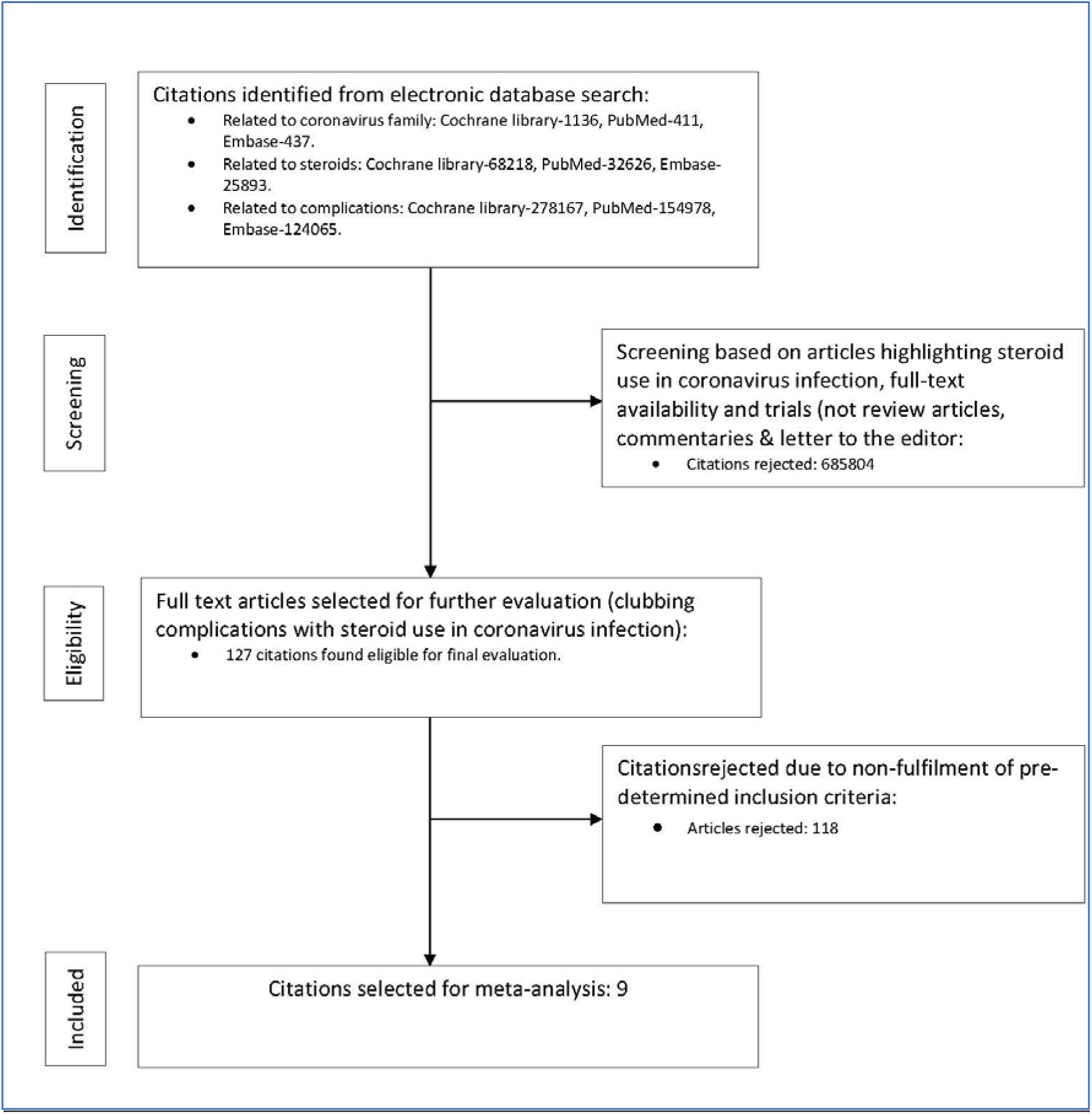
Study selection process

Due to a dearth of well-designed randomized prospective studies on the topic under consideration, a wide-angle search was conducted with the following inclusion criteria:

1. The trials must report a clear distinction between use and non-use of steroids as the two-comparator arms.
2. Studies must include viral infection belonging to the coronaviridae family only.
3. ARDS included as the complication of interest must be secondary to viral infection.
4. Although mortality was the prime outcome of interest, overall complications were included in the search to identify representation of mortality under divergent terminologies.
5. All patients in the active-treatment arm must have been treated with steroids. We excluded all data presented in the form of commentaries, review articles, or case reports. Studies reporting some patients getting steroids in the active arm were excluded from the analysis if this group was not represented as a separate entity. We also did not evaluate the studies in the pediatric age group.

### 2.2 Process of study selection

The study was conceptualized by KKG. The article screening process was performed in a cyclical manner to make the process more robust. In phase one, SG and JJM conducted the web search jointly with mutual consultations. Any dispute was resolved on the basis of a mutually acceptable consensus. The inclusion and exclusion criteria were devised by BS & KKG. Once the whole study selection process was completed, the team performed the same search in reverse order, with BS & KKG doing the web-based screening and JJM & SG formulating the selection criteria.

### 2.3 Study quality assessment

Quality of individual studies were assessed using the Cochrane collaboration tool using random sequence generation, allocation concealment, blinding of participants and personnel, blinding of outcomes assessment, incomplete outcome data, selective reporting and other bias as assessment attributes. Publication bias was assessed using funnel plots.

### 2.4 Statistical analysis

Analysis was conducted on a pooled patient population of 2,636 patients identified with confirmed coronavirus infection from nine citations (5,15-22), using the comprehensive meta-analysis software version 3, Biostat Inc., Englewood, NJ, USA. Heterogeneity was assessed using the Cochrane Q and Higgin’s I^2^ test, and publication bias was assessed by funnel plots. [1-9] Depending on the degree of heterogeneity (<45% low, 45–75% moderate and > 75% high) and study characteristics, a fixed or random effect model to assess the effect size was selected. Where relative-risk or odds-ratio were not reported, rate-ratio was calculated from the reported events using Medcalc statistical software, © 2020 MedCalc Software Ltd, Ostend, Belgium.

## 3.0 Results

We identified nine studies (three with SARS, one with MERS, and five with COVID-19) that met with the inclusion criteria (5,15-22). The details of the individual studies are noted in table 1. A total of 1179 patients (both critically & non-critically ill) were on steroids from a pooled patient population of 2880. Of these, 459 patients had ARDS diagnosed with a set of specific criteria. The meta-analysis initially compared the morality outcome in all patients (SARS, MERS & COVID-19; n=2880) who received CS with those who did not receive CS; subsequently, the mortality outcome in all patients with COVID-19 alone (n=1781) and in patients with COVID-19 alone and ARDS who received CS was compared to those who did not receive CS. Mortality data was analyzed by taking into consideration patients who were critically ill and/or in intensive care unit depending upon whether they were receiving CS or not.

**Table 1:** Characteristics of observational studies included in the meta-analysis. NR = not reported, ALI = Acute lung injury, MP = methylprednisolone, Dexa = dexamethasone, NR = not reported, n=total number of patients in respective studies, n1= total number of patients on corticosteroids (both critically & non-critically ill patients).

| Studies | Age | ARDS or ALI / total number of patients (n) | Steroid (Types) & n1 | Associated treatment | Early discharge (CS group) | La b/Radiological benefits (CS group) |
| --- | --- | --- | --- | --- | --- | --- |
| Lew et al (15) (SARS) | 51 (Median) | 45/199 | Pulse MP<br>38 | Ribavirin/oseltamivir<br>Broad-spectrum antibiotics | NR | NR |
| Zhao et al (16) (SARS) | 28.6± 10.3 | 36/190 | MP<br>120 | Ribavirin<br>Broad-spectrum antibiotics<br>Recombinant INF-α | NR | NR |
| Chen et al (17) (SARS) | 40.0±14.6 | 152/401 | MP (mostly)<br>Dexa (Minority)<br>370 | Ribavirin<br>Broad-spectrum antibiotics | OR 1.74,<br>95%CI: 1.02-2.96 | NR |
| Arabi et al (5) (MERS) | 57.9±17.2 | NR/309 | Hydrocortisone (most common), MP, prednisolone, Dexa<br>151 | Ribavirin/oseltamivir<br>Recombinant INF-α | NR | Delayed MERS coronavirus RNA clearance (adjusted hazard ratio, 0.35; 95% CI, 0.17–0.72; P = 0.005) |
| Wu et al (18) (COVID-19) | 51(Median) | 84/201 | MP<br>62 | Broad-spectrum antibiotics<br>Oseltamivir, ganciclovir, lopinavir/ritonavir<br>Recombinant INF- $\alpha$ | NR | NR |
| Zhou et al (19)<br>(COVID-19) | 46.0–67.0 | 59/191 | Corticosteroids<br>(?type)<br>57 | Lopinavir/ritonavir<br>IV Ig<br>Broad-spectrum<br>antibiotics | NR | NR |
| Guan et al (20)<br>(COVID-19) | 47 (Median) | 37/1099 | Systemic<br>Glucocorticoid<br>(?type)<br>204 | Broad-spectrum<br>antibiotics<br>Oseltamivir | NR | NR |
| Wang et al (21)<br>(COVID-19) | 48-64 | 46/46 | MP<br>26 | Broad-spectrum<br>antibiotics<br>Lopinavir/ritonavir<br>Thymosin | NR | Significantly better<br>improvement in SpO2<br>and faster resolution<br>of CT changes |
| Lu et al (22)<br>(COVID-19) | 53-71 | 87/244 | MP<br>Dexa<br>151 | Oseltamivir,<br>arbidol,<br>lopinavir/ritonavir,<br>ganciclovir, interferon-α | NR | NR |

### Group 1: Mortality in patients with SARS, MERS, and COVID-19 (Figure 2)

**Figure 2:**
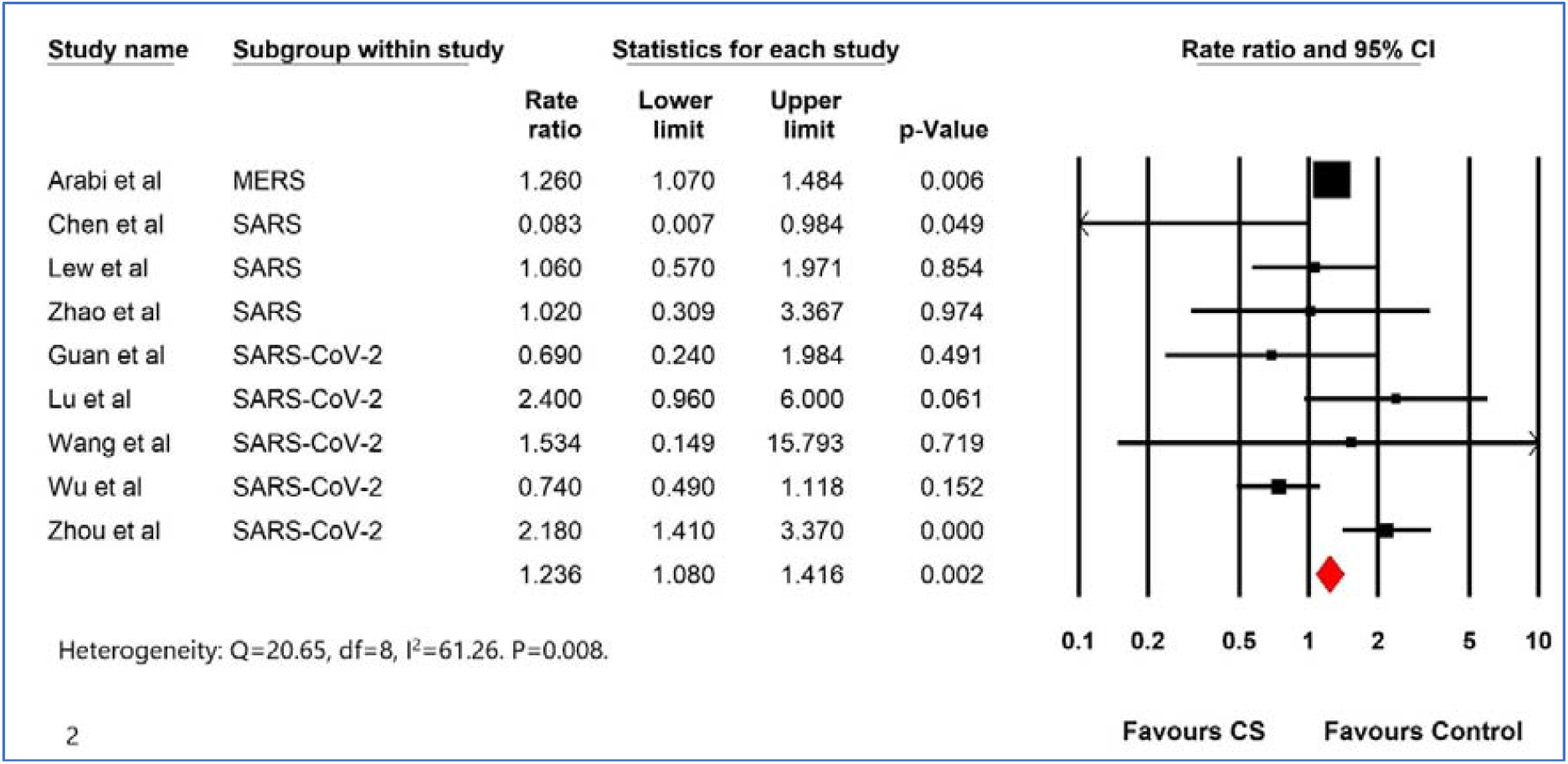
Forest plot comparing the mortality in patients with SARS, MERS, and COVID-19 who received systemic corticosteroids with those who did not receive CS. Black box indicates individual effect size and the red diamond-indicates the overall effect size.

The rate ratio for mortality was significantly higher in patients with SARS, MERS, and COVID 19 combined on CS when compared to those who did not receive CS (RR: 1.24, 95% CI: 1.08-1.42, I^2^: 61.26) (Figure 2). There was moderate heterogeneity.

### Group 2: Mortality in patients with COVID-19 (Figures 3a and 3b)

**Figure 3:**
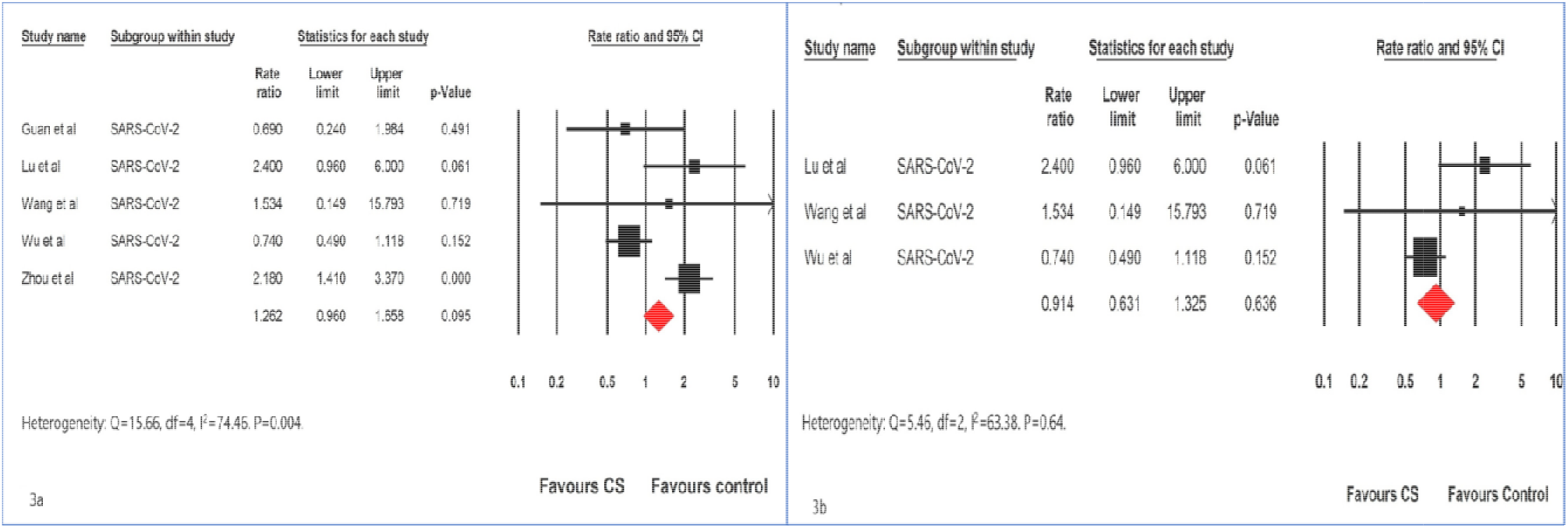
Forest plot comparing the mortality in critically ill patients with COVID-19 who received systemic corticoo with those who did not receive corticosteroids (a) all patients (b) patients with COVID-19 with ARDS.

There was no statistically significant difference in morality among the patients with COVID-19 who received CS when compared to those who did not receive CS (RR:1.26, 95% CI: 0.96-1.66, I : 74.46); there was moderate to high heterogeneity.

In two of the five studies included in the meta-analysis (Zhou et al and Guan et al), not all patients had ARDS (19,20). The individual rate ratios for mortality in the three studies (Lu et al, Wu et al and Wang et al) that recruited patients exclusively with ARDS/requirement for artificial ventilation were widely divergent (18, 21,22). Wu et al (18) had reported a reduction in mortality using methylprednisolone in patients with COVID-19 with ARDS (HR: 0.38, 95%CI, 0.20 – 0.72); however, as analyzed here, the rate ratio of mortality (RR: 0.74; 95% CI: 0.49-1.11), based on the events at a point in time, was not significant. Meta-analysis of these three studies (18,21,22) did not show statistically significant difference in the mortality between the patients who received CS when compared to those who did not (RR: 0.91, 95% CI: 0.63-1,32, I : 63.38)

## Discussion

The pathogenic mechanism of infection and toxicity for SARS and SARS-CoV-2 is a result of an interplay between the viral receptor, viral replication, and host immune response (23). This results in a systemic inflammatory response, increase in pulmonary vascular permeability, and a cytokine storm, culminating into ARDS (23).

There is no specific pharmacological treatment for ARDS. Steroids, with their potent anti-inflammatory and anti-fibrotic properties, have been tried in ARDS with mixed results. In 2017, the Society of Critical Care Medicine (SCCM) together with the European Society of Intensive Care Medicine (ESICM) reviewed nine RCTs and based on moderate quality evidence, recommended the use of steroids in patients with moderate-to-severe ARDS (24). There is also some uncertainty regarding the use of CS in septic shock stemming from the differences in the results of the four large RCTs (25-28); although all four trials showed benefits in haemodynamic status, only two showed survival benefit (25,28). The 2017 SCCM/ESICM guidelines recommend use of low dose intravenous (iv) hydrocortisone (<400 mg/day) for at least 3 days or longer in adult patients with septic shock not responding to fluid and moderate-to high-dose vasopressor therapy (> 0.1 ug/kg/min of norepinephrine)(24).

The effect of use of CS in viral pneumonias of mixed etiology remains uncertain. Whilst a recent meta-analysis showed an association between CS use and an increase in mortality in patients with viral pneumonias, the effect of use of CS in patients with coronavirus infections in other studies was unclear (29). Although a recent Cochrane review on management of ARDS of mixed etiology suggested that CS treatment reduced mortality (RR, 0.75; 95% CI, 0.59 to 0.95) (30), this cannot be generalized to patients with SARS-CoV-2 infection as these trials were not focused on ARDS secondary to viral etiology in general, and SARS-CoV2 infection in particular.

We found a statistically significant increase in mortality with the use of CS among 2880 critically ill patients with coronavirus (SARS-CoV, MERS-CoV, SARS-CoV2) infections. This was mainly driven by the single large study involving patients with MERS (5), which alone had more number of events than the rest of the studies combined together. Genetically, the similarity of SARS – Cov-2 to SARS is about 79%, and to MERS is about 50% hence SARS – CoV-2 is more phylogenetically related to SARS than to MERS (32). While SARS-CoV-2 and SARS share the same functional human cell receptor, the angiotensin-converting enzyme 2 (ACE2), MERS coronavirus uses dipeptidyl peptidase 4 (DPP4) to enter host cells (33). MERS affected patients have a higher mortality with higher rates of acute kidney injury than SARS-CoV-2 infection, indicating some differences in the pathogenesis (34). Moreover, there was moderate heterogeneity in these observational studies concerning choice of patients, type, doses, and timing of steroid administration, and the presence or absence of ARDS, all with a potential for a significant impact on the outcome.

We did not find any statistically significant difference in mortality with the use of CS in the 1781 critically ill patients with SARS-CoV-2 infection. In a recent multicenter, randomized controlled trial in patients with established moderate-to-severe ARDS of varying etiology, the use of intravenous dexamethasone resulted in less number of deaths in the dexamethasone arm at 60 days (between-group difference –15·3% [–25·9 to –4·9]; p=0·0047) (31). Moreover, other parameters including ventilator free days at 28 days, duration of mechanical ventilation in ICU survivors, duration of mechanical ventilation at day 60 all favored CS use in this study (31). We therefore analyzed the effect of use of CS in critically ill patients with COVID-19 with ARDS in the three studies (18,21,22) that had reported on this. In absence of uniform reporting of all outcomes of interest, we confined ourselves to the effect of use of CS on mortality. Similar to our finding for the whole cohort of critically ill patients with COVID-19, we did not find any statistically significant improvement in mortality among the 499 of 1781 patients with COVID-19 with ARDS. Individually, the effect of use of CS among critically ill patients with COVID-19 in these five observational studies (18-22) was very varied. In the retrospective cohort study by Wu et al (18) involving 201 patients with COVID-19, use of methylprednisolone reduced the risk of death among the 84 patients with ARDS [23/50(46%) who received CS died, and 21/34 (61.8%) who did not receive CS died; (hazard ratio, 0.38, 95%CI, 0.20–0.72)]. However, since events at different quintiles were not reported, and since all the other studies reported relative risk/odds ratio, the risk ratio was calculated and used for the study by Wu et (18) al in order to maintain a standardized pattern of reporting. Zhou et al (19), studied 191 patients to explore the risk factors associated with in-hospital death of SARS-CoV-2 patients and found that 23% of patients who survived received CS while 48% of non survivors did not receive CS. Wang et al (21) reported 46 patients of COVID-19 with severe pneumonia and found that patients who received intravenous methylprednisolone 1-2 mg/kg/d for 5-7 days (26 patients) had significantly lesser number of days for body temp to normalize (2.6 vs 5.29, p=0.10), shorter interval of using supplemental oxygen therapy (8.2 v/s 13.5 days, p < 0.001) and had better degree of absorption on chest CT. There were only 3 deaths, two of them in the CS group. Guan et al (20) extracted data of 1099 patients with COVID-19; out of a total of 15 deaths, five occurred among the 204 patients who received CS. However, CS was given in a higher percentage of patients with severe disease when compared to those with non-severe disease (44.5% vs. 13.7%), which can explain the higher mortality in the CS group. Lu et al (22) evaluated the effects of CS treatment on 244 critically ill patients with COVID-19. In the case control group, the calculated rate ratio for mortality was 2.4 (95% CI; 0.96 - 6.00, p = 0.061).

There are a number of limitations in our meta-analysis. Firstly, the studies included for analysis used different primary and secondary criteria as aims for their respective studies resulting in a moderate degree of heterogeneity. Secondly, there were significant differences in the types of steroids used in the individual studies. This could have an impact on the final analysis. Thirdly, we are in the midst of a pandemic, and new data is being accrued constantly, which might impact our results. Fourthly, we have used rate ratio over hazard ratio to report mortality outcomes as it is a better index for assessing mortality over time. Majority of the trials included in the meta-analysis did not report a hazard ratio or mortality events over a period of time. We therefore took the cumulative events as reported at the end of the study period, ignoring time frame, for calculations. [Supplementary table 1]

The strength of our meta-analysis lies in the fact that in an evolving infectious disease pandemic with acute consequences, the best way to increase the predictability of the question in focus is to perform pooled analysis. By pooling data and performing a systematic review and meta-analysis we have circumvented the problems associated with individual studies that have very divergent results as noted above; this shall help guide physicians till further information becomes available in the future. Secondly, to bring in some degree of uniformity, we created a stringent inclusion criterion. Since, the parameters assessed and the outcomes analyzed varied enormously between the studies, we decided to focus on one of the most important parameters assessed in the critical care unit i.e. mortality. The associated risks and benefits were not ignored, but analyzed on a case to case basis. As such, the clinical and radiological improvement with corticosteroid as reported by Wang et al (21) were not taken into consideration as they were not uniformly reported in the four other studies analyzed.

. In conclusion, CS can be given in critically ill patients with COVID-19 with CIRCI and considered in those with ARDS, strengthened by the fact that our meta-analysis does not show an increase in mortality in the critically ill patients with COVID-19 who received CS. Our meta-analysis strengthens the recommendation of the Surviving Sepsis Campaign COVID-19 panel of using low doses of CS for a short duration in the critically ill patients with COVID-19 with ARDS.

## Conclusion

To our knowledge this is the first meta-analysis of CS use in patients with coronavirus infections in general, and SARS-CoV-2 in particular. Although limited by confounding factors typical of retrospective studies, this meta-analysis indicates lack of harm with use of systemic CS in critically ill patients with COVID-19 with ARDS. We believe that this meta-analysis may help physicians and intensivists to consider CS therapy in deserving critically ill patients with SARS-CoV-2 infection.

## Data Availability

All data used are in public domain.

